# Number of Austrian SARS-CoV-2 infections in the 2024/2025 season: Analysis of national wastewater data

**DOI:** 10.1101/2025.05.05.25327005

**Authors:** Uwe Riedmann, Wolfgang Rauch, Hannes Schenk, Herbert Oberacher, John PA Ioannidis, Stefan Pilz

## Abstract

Adaptation of SARS-CoV-2 policies requires information of contemporary infection trends. Based on wastewater data, we estimate 2.5 million new infections in Austria between April 1, 2024, and March 31, 2025, with peak in early October. This indicates the earliest annual fall/winter peak and the lowest annual infections since 2020.

## INTRODUCTION

To create and update health policies regarding SARS-CoV-2, it is imperative to have a grasp on the current disease burden. That includes knowledge on the number of infected, and overall infection trends. As the SARS-CoV-2 infections are no longer actively tracked, we need to utilize indirect measures such as estimation from wastewater data.

Raw wastewater data can give us insight in trends and changes in infection frequencies across different years, and estimation of infections further provide scaling, enabling comparisons to influenza and other diseases. Such estimations have previously been performed for Austria up to May 2024.^1–3^

Here we used a previously published model on recent national wastewater data to estimate the number of all SARS-CoV-2 infections that occurred in Austria from April 2024 1, to March 31, 2025.^1^

## METHODS

### Study data and analysis

We conducted a retrospective estimation of total SARS-CoV-2 infections in the entire population of Austria from April 1, 2024 to March 31, 2025, based on wastewater monitoring data.^1,2,4^ We chose this time period as the nadir of estimated active infections during 2024 occurred in late March. Wastewater data from the Austrian SARS-CoV-2 wastewater monitoring initiative was provided by the Austrian Federal Ministry of Labour, Social Affairs, Health, Care and Consumer Protection for the period from November 2022, to April 21, 2025. Since 2023, the dataset covers the unchanged catchment area of approximately 58% of the Austrian population. Data before December 2022 was based on a publication by Rauch and colleagues.^1^ We additionally provide estimates for the calendar years.

The study was approved by the ethics committee at the Medical University of Graz (no. 33-144 ex 20/21). The statistical analyses and simulations were conducted using R (version 4.4.2).^5^

### Infection estimation from wastewater data

For the estimation of total SARS-CoV-2 infections we applied our previously published approach, that estimated active infected and new daily infections in Austria based on wastewater data from May 2020 to May 2024.^1,3^ The wastewater data was filtered, outlier corrected, averaged and interpolated before the model estimation.^2,6–8^ We then applied the wastewater model from Rauch et al.^1,4^ and scaled parameter estimates documented in Riedmann et al.^3^, to estimate daily active infections. A backcasting algorithm was applied to estimate daily new infections from daily active infections (see Supplements). The monitoring data as well as the pre-processing and normalization methodology is presented in the supplementary methods, and described in detail in previous publications.^1–4,6–8^

## RESULTS

### Infection estimation from wastewater data

Between April 1, 2024, and March 31, 2025, estimated active infected individuals fluctuated between 337,000 at its peak on September 27-29 and 8,800 at the lowest on April 1-4, 2024 (Figure 1). Within this period, we estimated a total of approximately 2.5 million new infections (Figure 2).

**Figure 1:**
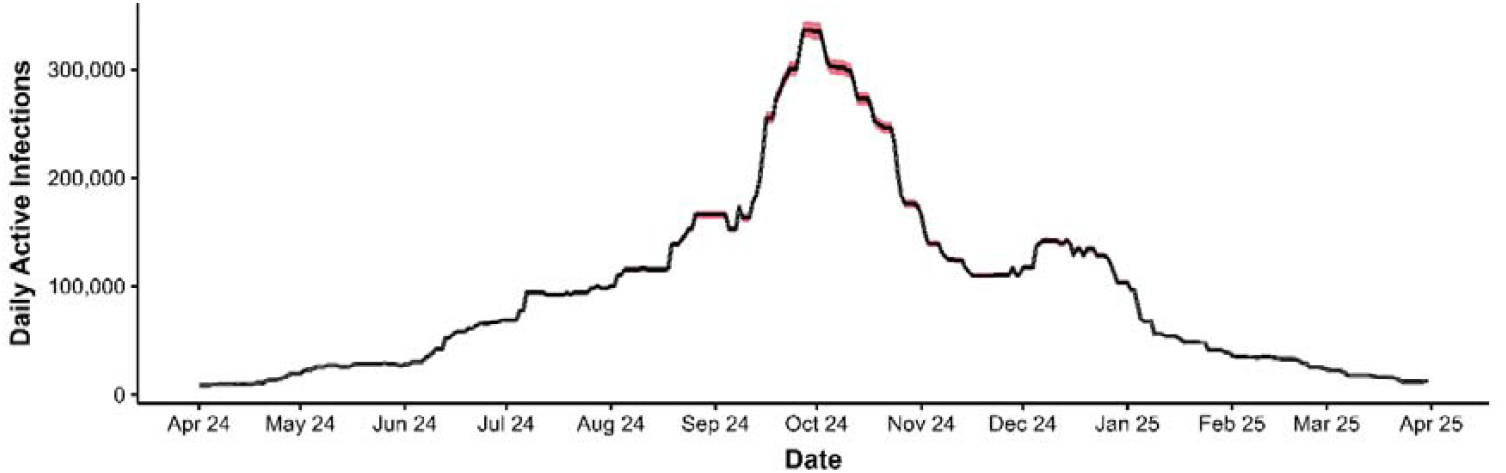
Active daily infection estimates. Red area indicates confidence intervals (CI) of the model parameter.^1^ The peak is on 27-29, September 2024. For the calendar years 2020, 2021, 2022, and 2023, the fall/winter peaks had occurred in November 20, November 24, October 8, and December 11-13, respectively, while 2022 had also an even higher spring peak (March 22, 2022) (Figure S1a). The CIs indicate uncertainty in the originally estimated correction factor (shedding and loss; see Supplements).^1^ They do not incorporate potential changes in shedding intensity and shedding duration since 2022 and thus result in relatively narrow CIs.

**Figure 2:**
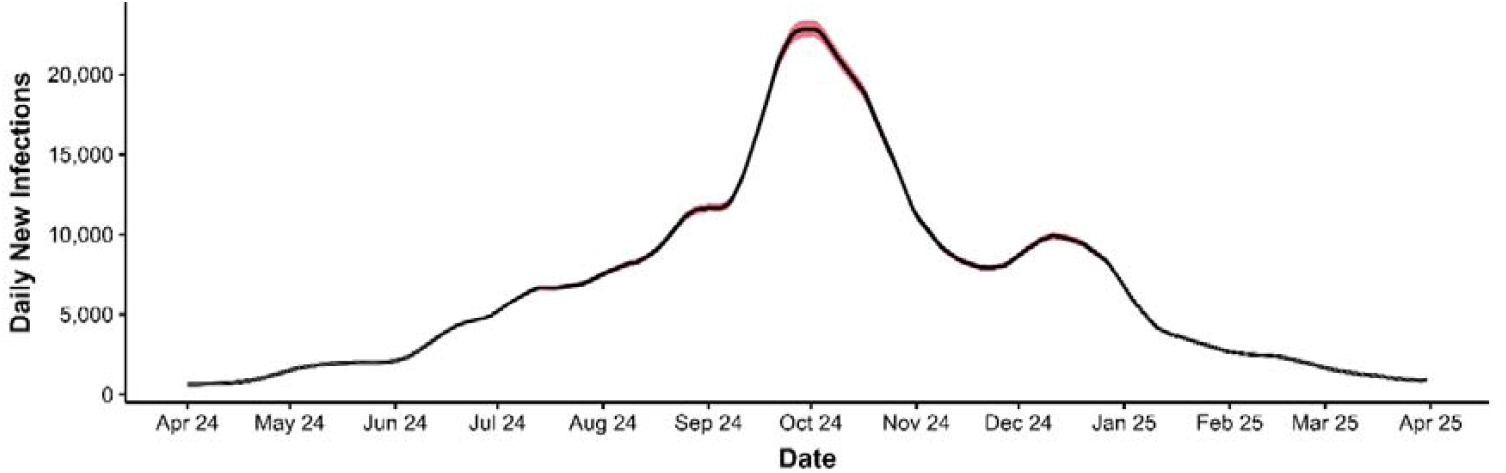
New daily infection estimates. Red area indicates confidence intervals of the model parameter.^1^ The peak is on 2, October 2024. For the calendar years 2020, 2021, 2022, and 2023, the fall/winter peaks had occurred in November 17, November 24, October 2, and December 13, respectively (Figure S1b). Note that slight shifts in peaks compared to the active daily infected estimates are due to smoothing during the backcasting algorithm. Similar caveats exist about the narrow confidence intervals as for Figure 1.

In previous years 1.8 million, 4.5 million, 5.5 million and 3.3 million estimated infections occurred starting in April 2020, 2021, 2022 and 2023, respectively (Figure S1). Estimated infections of calendar years were at 2.8 million for 2024 as well as 1.1 million, 1.9 million, 7.4 million and 4.2 million for 2020, 2021, 2022 and 2023 respectively (Figure S1). Note that estimates for 2020 were only available after May 19th.^3^

## DISCUSSION

We found that the SARS-CoV-2 infection rates still fluctuated markedly across seasons, but the primary fall/winter wave was much earlier in 2024 than in previous years. After an early peak in September, infections stayed at a moderate to low level throughout the winter and have plummeted in spring 2025. Overall, 2.5 million estimated infections occurred between April 1, 2024, and March 31, 2025 (28% of the population size). This represents a decrease in infections of about a quarter compared to the same time period 2023/2024.^3^

Wastewater data allow calculation of “active” infections that reflect viral shedding. Hence, one can calculate new daily infections (and total infections over a given period) based on an assumed estimate of the duration of viral shedding. The decrease in active infections is assumed to reflect fewer daily infections but it can also be affected by a change in the duration in viral shedding. Increased immune protection in the general population may be responsible for the decrease in infections.^3^ Alternatively, higher immunity may have reduced the severity of the disease and duration of viral shedding.^9,10^ E.g. if duration of viral shedding has decreased to 10 days instead of 14, then the estimated number of infected in that last year is 3.5 million instead of 2.5 million.

Overall, substantial parts of the population are infected every year, possibly contributing to enhanced immunological protection on a population level.^3^ Whether these continuous immunizations by repeated SARS-CoV-2 infections obviate the future need for SARS-CoV-2 routine testing and vaccinations warrants further studies.

The findings are likely to be representative in scale for a majority of western countries in the northern hemisphere, as Austria is comparable to most western European countries in its social system and healthcare system.^3^

The results are subject to several methodological limitations, as discussed in previous publications, and should be interpreted as rough estimates.^1,3^ For the calculations we used an estimated population size of 9.02 million. This estimate is based on previous, mid pandemic population statistics. Since then, Austria’s population has increased by about 2% (January 2025: 9.2 million).^11^ Using this higher base population increased estimated new infections by about 50,000. We deemed this difference neglectable and decided to keep the previous population size to ease comparability between years.

As fatality rates were already very low by 2022,^12^ in the endemic phase policy makers, physicians and high-risk individuals should be aware of the ongoing seasonal SARS-CoV-2 infection waves and their variability in timing. Future studies may also link SARS-CoV-2 infection data as provided by our study to mortality, hospitalization and health utilization data in order to assess cost-effectiveness calculations on COVID-19 policies.

## Data Availability

The data that support the findings of this study are available upon request with approval needed from the Austrian Federal Ministry of Labour, Social Affairs, Health, Care and Consumer Protection.

## Contributors

UR and SP conceptualized the study. UR and SP wrote the original draft of the manuscript. SP acquired funding. UR and HS wrote software. HO curated data. UR performed the formal analyses and handled visualization. SP administered the project. All authors contributed to writing, reviewing, and editing the manuscript, and approved the final version before submission.

## Declaration of interests

The authors declare no conflict of interests.

## Funding

The study was founded by the Austrian Science Fund (FWF) KLI 1188.

For this study, data from the National SARS-CoV-2 Wastewater Monitoring Program was used. This program is financed by the Austrian federal Ministry of Labour, Social Affairs, Health, Care and Consumer Protection who has the sole right of use for the data.

## Acknowledgements

The authors thank all persons and organizations involved in data collection.

## Notes

### Competing Interest Statement

The authors have declared no competing interest.

### Author Declarations

The study was approved by the ethics committee at the Medical University of Graz (no. 33-144 ex 20/21).

