## Supplements for "Number of Austrian SARS-CoV-2 infections in the 2024/2025 season: Analysis of national wastewater data"

### **Supplementary Methods**

#### **Wastewater** **Model**

##### **Data** **Description**

The dataset on wastewater data provided by the Austrian Federal Ministry of Labour, Social Affairs, Health, Care and Consumer Protection spanned the period from November, 2022, to April 16, 2025. It included 48 wastewater treatment plants (WWTP) from 2023 onward (24 before), with 11,195 measurements total. These treatment plants cover approximately 58% of the Austrian population. Wastewater samples were collected twice a week.

Viral concentration is determined and processed as previously explained [1]. Additionally, different hydrochemical parameters were used for characterizing the catchment population of the monitored wastewater treatment plants. For the chemical oxygen demand (COD) estimates, per capita equivalents were calculated using 120 g/d/person. Ammonia-nitrogen (NH4-N) estimates were calculated using 8.0 g/d/person. Total-nitrogen (Ntot) estimates were calculated using 11.0 g/d/person [2].

##### **Data** **Preprocessing**

For a detailed description of preprocessing see previous publications [1,3].
To compensate for inherent measurement noise, Rauch et al. [4] suggest the approach to exclude outliers if the flow volume (Q) exceeds the 90 percentile of the long term recorded inflow data of a WWTP (needs at least a year of data points).

Estimates are normalized based on population-size markers, to compensate for population fluctuations within a catchment area. Following Arabzadeh et al. [2], we used NH4-N prioritised over COD and Ntot. We computed the daily weighted averages of viral load levels per federal state. The weights correspond to the design capacity of the respective WWTPs, prioritising large plants over smaller ones. The design capacity of each WWTP is a parameter, that serves as a weighting factor when computing the weighted average of multiple measurements in spatial aggregation. In principle, the preferred weighting factor is the exact catchment population. However, this information is currently unavailable to us.

This results in a scattered time-series from WBE measurements that are not gapless on a daily basis. We used upsampling to get daily estimates by linearly interpolating gaps before applying data smoothing [5]. Lastly, data filtering techniques are applied to reduce the signal noise and provide a mechanism to obtain the underlying information of the signal.

##### **Model** **Description**

The measured virus load at the monitoring point is related to the population drained with the sewer system:

$$L_{virus}=\frac{c_{virus}*Q}{N}$$

Where L_virus_ is the population normalized virus load in gene copies/**p**erson/**d**ay, Q is the flow volume in Liters/day, c_virus_ is the virus concentration in the sample in copies/Liter and N is the catchment population.

Under the assumption that each infected person is shedding a certain load of gene copies per day (L_shed_ in gene copies/person/day) into the sewer system and additionally introducing a general loss term f_loss_ we get:

$$I\left( t+t_{lead} \right)=\frac{L_{virus}(t)\times N}{L_{shed}\times f_{loss}}=\frac{L_{virus}(t)\times N}{L_{corr}}$$

Where *I* is the number of infected individuals in the watershed, t_lead_ is the time lead and f_loss_ is a dimensionless loss factor.

We used t_lead_ = 0 and L_corr_ = 10^10.090^ akin to Riedmann et al (2024)[6]. The population was set to *N* = 9.02 × 10^6^ [7]. As these values represent currently infected, we needed to apply a backcasting algorithm to estimate daily new infections.

##### **Backcasting**

The backcasting methodology used here is designed to estimate daily infections based on daily active cases. The key assumption is that an infection lasts 14 days on average.

We first smoothed the estimated undocumented daily infection counts via 14 days centred moving average. The core of the backcasting process involves iteratively refining the infection level estimates, which were initially based on the smoothed data. Potential estimation errors from variability in counting method was addressed by setting negative testing values to 0. Lastly, after refining the estimates, a secondary smoothing step was applied to the calculated infection values.

In accordance with a previous publication we up scaled the estimates by 25% to align with seroprevalence data [6].

### **Supplementary Results**

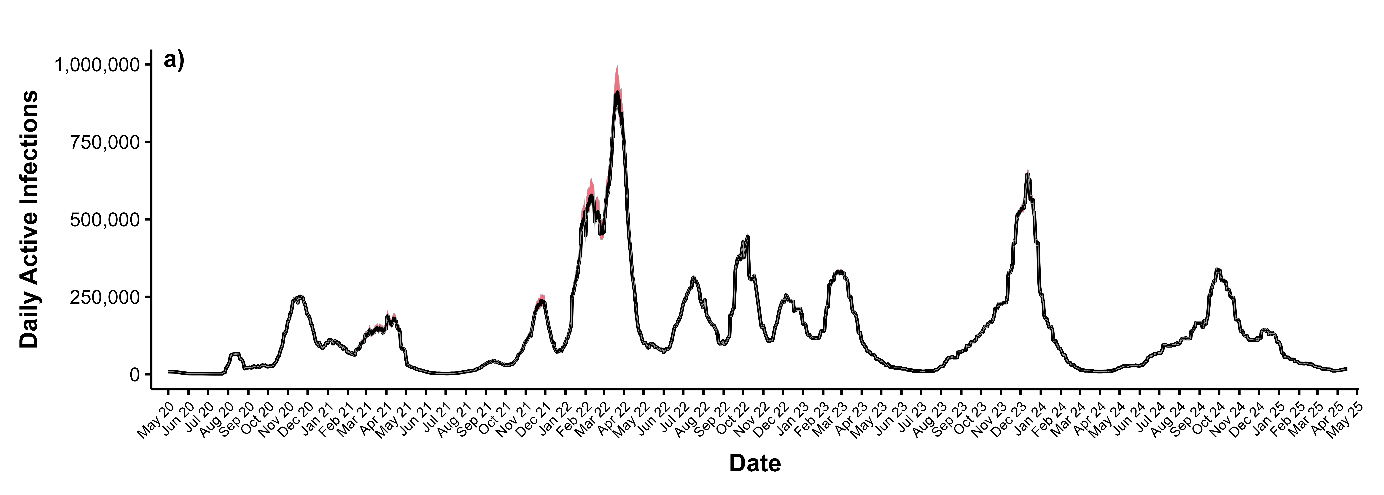

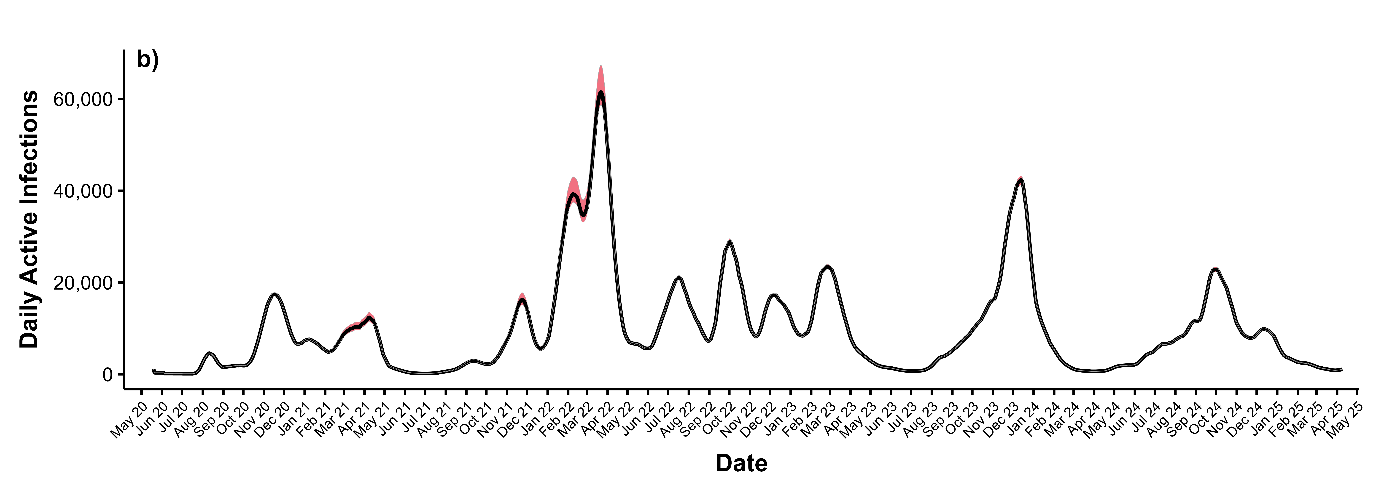

**Figure S1:** Estimates of active infection (a) and daily new infections (b) between May 2020 and April 2025

### **Supplementary Discussion**

#### **Limitations**

Wastewater

There are measurement errors in the determination of the hydrochemical parameter concentration. While outliers are addressed as mentioned in the pre-processing steps, smaller errors, akin to noise, cannot be avoid.

The shedding rate varies from person to person. The average shedding rate is also not constant. The immunization rate and the virus variant are potential factors that can also influence the average shedding rate.
